# Calibrated and Interpretable Machine Learning for ICU Mortality Prediction Using First 24-Hour Clinical Data

**DOI:** 10.64898/2026.05.30.26354524

**Authors:** Abdallah Alsammani, Merasia Johnson, Jessica Elrefaei

## Abstract

**Objective:** To develop, calibrate, and interpret machine learning models for predicting in-hospital mortality among intensive care unit (ICU) patients using clinical data from the first 24 hours of admission.

**Methods:** We analyzed 53,866 adult ICU admissions from MIMIC-IV (v2.2), including 5,787 in-hospital deaths (10.7%). An enhanced feature-engineering pipeline generated 88 laboratory features capturing distributional characteristics, temporal trends, and measurement frequency. Five classifiers were evaluated: *𝓁*_2_-regularized logistic regression, random forest, XGBoost, LightGBM, and a calibrated soft-voting ensemble. Models were developed using a stratified 64:8:8:20 split for training, validation and hyperparameter tuning, calibration, and testing. Performance was assessed on a held-out test set (*n* = 10,774) using AUROC, AUPRC, Brier score, calibration analysis, decision curve analysis (DCA), and SHAP-based interpretation.

**Results:** The calibrated ensemble achieved the best overall performance (AUROC 0.856, 95% CI 0.846–0.867; AUPRC 0.449, 95% CI 0.418–0.480) with a Brier score of 0.078. XGBoost (AUROC 0.856; AUPRC 0.435) and LightGBM (AUROC 0.854; AUPRC 0.436) performed comparably to the ensemble and significantly outperformed logistic regression (AUROC 0.823; AUPRC 0.376), yielding absolute AUROC improvements of approximately 0.031–0.033 (*p <* 0.001). Calibration reduced Brier scores by 42% for XGBoost (0.134 to 0.078) and 50% for LightGBM (0.151 to 0.076). Decision curve analysis demonstrated consistent net benefit across the 5%–20% risk-threshold range. Key predictors included age, blood urea nitrogen, ICU subtype, measurement frequency, and lactate-related features, with consistent performance across ICU subtypes (AUROC *>* 0.79).

**Conclusion:** A calibrated and interpretable machine learning framework using early ICU data provides accurate and clinically actionable mortality risk estimates. By integrating trajectory-aware feature engineering, probabilistic calibration, and decision-analytic evaluation, this approach advances ICU mortality prediction toward reliable clinical decision support.

## 1 Introduction

Predicting in-hospital mortality in the intensive care unit (ICU) is a central clinical priority, with implications for resource allocation, escalation of care, and goals-of-care discussions. Conventional severity scores such as APACHE II [1] and the Sequential Organ Failure Assessment (SOFA) score [2] aggregate physiological variables into composite indices using hand-crafted rules derived from regression analyses of relatively small, single-center cohorts. Their discrimination tends to degrade outside the original development setting [3], and their manual computation introduces operational friction in time-pressured environments.

The release of large critical care databases has broadened the empirical landscape of ICU mortality prediction research. MIMIC-IV [4] provides de-identified electronic health records from more than a decade of ICU admissions at a single academic medical center and serves as a rigorous benchmark for predictive modeling. Traditional machine learning methods remain well-suited to this setting because they are computationally efficient, naturally compatible with heterogeneous tabular clinical variables, and can capture nonlinear predictor relationships without requiring high-dimensional sequential modeling [5]. Gradient-boosted tree ensembles, in particular, have shown strong performance across clinical tabular benchmarks [6, 7], while SHAP (SHapley Additive exPlanations) provides a principled framework for patient-level interpretability and model transparency [8, 9]. Accordingly, this study emphasizes practical, interpretable, traditional machine learning approaches for ICU mortality prediction.

Three methodological gaps persist in the published ICU mortality literature using traditional machine learning. First, many existing approaches rely primarily on static aggregation of laboratory and physiologic variables using summary statistics such as minimum, maximum, and mean values, without explicitly representing short-term clinical trajectories or measurement intensity, both of which may contain clinically relevant prognostic information [12, 24, 30, 31]. Second, probability calibration and decision-analytic evaluation are frequently omitted, despite their importance for threshold-based clinical use [18]. Third, the comparative behavior of modern gradient-boosted tree architectures, together with calibrated ensembling, has not been systematically reported on MIMIC-IV.

The present study addresses these gaps through three contributions. We introduce an enhanced feature engineering pipeline that extracts eight statistics per laboratory analyte (including linear trend and measurement count), yielding 88 laboratory features compared with the 33 features in standard pipelines. We compare five classifiers, including a calibrated soft-voting ensemble, with rigorous DeLong-based statistical testing. We provide a comprehensive evaluation framework that combines discrimination, calibration before and after isotonic regression, decision curve analysis, ICU-subtype subgroup discrimination, and SHAP-based interpretation.

### Novelty and Contributions

This study introduces a unified and clinically grounded framework for ICU mortality prediction that differs from prior work in several key aspects. First, we incorporate both temporal trend features and measurement intensity (count features), capturing dynamic physiological trajectories and clinical attention patterns that are typically ignored in standard pipelines. Second, we provide a rigorous evaluation of probability calibration using isotonic regression and demonstrate its impact on model performance and clinical utility. Third, we combine discrimination, calibration, and decision curve analysis within a single framework to explicitly assess clinical deployment readiness. Finally, we identify and characterize a previously underreported failure mode of random forest models under post-hoc isotonic calibration in imbalanced clinical settings, providing both empirical evidence and theoretical explanation.

## 2 Related Work

Clinical severity scores such as APACHE II [1] and SOFA [2] remain widely used for ICU risk stratification because they are standardized, interpretable, and straightforward to implement at the bedside. However, external validation studies have reported only moderate discrimination for APACHE II, with AUROC values between 0.74 and 0.82 and greater performance degradation under case-mix shift [3]. These scores rely on fixed predictor sets and prespecified aggregation rules, limiting their ability to capture nonlinear relationships, higher-order interactions, and institution-specific patterns in heterogeneous ICU data.

The public release of large critical care databases, beginning with MIMIC-III [13] and followed by MIMIC-IV [4], enabled more systematic evaluation of machine learning methods for ICU mortality prediction. Much of this work has relied on traditional tabular machine learning rather than sequential deep learning models. Pirracchio and colleagues demonstrated that super-learner ensembles outperformed APACHE II across two independent ICU cohorts [3]. Awad and colleagues used early ICU measurements and reported strong mortality discrimination with ensemble learning methods [12]. Zhu et al. developed machine learning models to predict mortality among mechanically ventilated ICU patients using first-day demographic, comorbidity, severity score, vital signs, and laboratory data from MIMIC-III, reporting strong predictive performance [32]. Building on this body of work, the present study focuses on practical, interpretable tabular machine learning methods well-suited for clinical deployment and decision support. More recent studies have similarly evaluated tree-based and gradient-boosted models such as random forests, XGBoost, LightGBM, and CatBoost for ICU mortality prediction using large public databases, including MIMIC and eICU [30, 31]. These studies support the practical value of traditional machine learning for critical care risk prediction, particularly when working with heterogeneous tabular clinical features.

In parallel, deep learning studies have modeled ICU time series directly, including benchmark tasks for in-hospital mortality, decompensation, length of stay, and phenotype classification [11]. However, the advantages of deep learning are less consistent in tabular settings, where tree-based ensembles often remain highly competitive while requiring less specialized architecture design and offering practical computational advantages [29]. The present study therefore focuses on deployable tabular machine learning methods rather than high-dimensional sequential deep learning models.

Despite progress in traditional machine learning for ICU mortality prediction, several limitations remain. First, many studies represent the early ICU period using static summary features such as minimum, maximum, and mean laboratory or vital-sign values. While clinically useful, this representation may discard short-term temporal changes, measurement frequency, and instability patterns that can carry prognostic information. Second, model evaluation often emphasizes discrimination, typically AUROC, while giving less attention to probability calibration and decision-analytic utility, both of which are important for threshold-based clinical deployment [18]. Third, although modern gradient-boosted tree architectures are widely used in tabular prediction tasks, their comparative performance, calibration behavior, interpretability, and ensemble combinations have not been systematically evaluated on MIMIC-IV for ICU mortality prediction.

The present study addresses these gaps by focusing on practical, traditional machine learning methods for ICU mortality prediction using the MIMIC-IV dataset. We compare five tabular classifiers: *𝓁*_2_-regularized logistic regression, random forest, XGBoost, LightGBM, and a calibrated soft-voting ensemble. In addition to conventional static summaries, we incorporate engineered features representing short-term clinical trends and measurement intensity. We further evaluate models using discrimination, calibration, SHAP-based interpretability, and decision-curve analysis to assess both predictive performance and potential clinical utility.

## 3 Materials and Methods

### 3.1 Ethical considerations and data access

This study used the publicly available MIMIC-IV database [4], distributed through PhysioNet [21] under a credentialed data-use agreement. Both authors completed the required CITI Data or Specimens Only Research training and signed the PhysioNet Credentialed Health Data Use Agreement before data access. Because the database contains only retrospective, fully de-identified records and the analysis posed no risk to patients, the study was exempt from institutional review board approval under 45 CFR 46.104(d)(4) and the terms of the PhysioNet data-use agreement.

### 3.2 Study population and outcome

MIMIC-IV (v2.2) contains de-identified clinical records from adult patients admitted to the ICU at Beth Israel Deaconess Medical Center, Boston, between 2008 and 2019. Inclusion criteria were: (i) age ≥ 18 years at the time of ICU admission, and (ii) at least one valid laboratory measurement within the first 24 hours of ICU admission. Exclusion criteria were: (i) ICU stays shorter than four hours (insufficient observation window), and (ii) for patients with multiple ICU admissions during the same or different hospitalizations, all non-index admissions were excluded to avoid within-patient correlation. The final analytic cohort comprised 53,866 admissions. The primary outcome was in-hospital mortality, defined by the MIMIC-IV admissions table field `hospital_expire_flag`.

### 3.3 Enhanced feature engineering

All features were derived exclusively from data available within the first 24 hours of ICU admission. Eleven laboratory analytes were selected on the basis of three criteria: (i) established prognostic value in published ICU severity scores and clinical practice (lactate, BUN, creatinine, electrolytes, hepatic enzymes, hematology indices) [1, 2, 25]; (ii) availability in ≥ 90%of admissions within the first 24-hour window in MIMIC-IV, ensuring stable feature extraction without excessive imputation; and (iii) coverage of the major organ systems implicated in critical illness, namely circulatory (lactate), renal (BUN, creatinine), electrolyte and acid–base (sodium, potassium, chloride), metabolic (glucose), hepatic (alanine aminotransferase, ALT), and hematologic (white blood cell count, hemoglobin, platelets). The corresponding MIMIC-IV laboratory item identifiers were: lactate (50813), BUN (51006), creatinine (50912), sodium (50983), potassium (50971), chloride (50902), glucose (50931), white blood cell count (51301), hemoglobin (51221), ALT (50878), and platelets (51265). Plausibility bounds were applied to remove implausible values attributable to transcription errors or unit-conversion artifacts before aggregation.

Vital signs and comorbidities were intentionally excluded from the present analysis to isolate the predictive contribution of laboratory data and to ensure consistent feature availability across admissions. Vital signs in MIMIC-IV are recorded at heterogeneous frequencies and through different chart-event identifiers across ICU subtypes, which complicates standardized feature extraction without dedicated time-series modeling. Comorbidity coding (ICD-9/ICD-10) is administrative and is finalized at discharge rather than at admission; its use as an admission-time predictor would constitute information leakage. Both data sources are reserved for future work, as discussed in Section 6.

For each analyte, eight statistics were computed within the 24-hour window, producing 11 *×* 8 = 88 laboratory features: minimum, maximum, mean, range (maximum minus minimum), count (number of measurements), first value, last value, and linear trend slope. Letting *x*_first_ and *x*_last_ denote the first and last measured values of an analyte and Δ*t* the elapsed time in hours between them, the trend feature is defined as

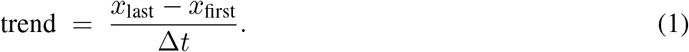

A linear approximation was chosen for two reasons. First, within a 24-hour window, most analytes are sampled too sparsely (median two to three measurements) to support higher-order parameterizations without overfitting at the patient level. Second, the sign and magnitude of a linear slope provide a stable summary of directional change that is robust to the precise sampling time and to small fluctuations between intermediate values. The count feature serves as a proxy for clinical attention intensity, reflecting nursing and physician concern about a particular organ system through the frequency of laboratory ordering.

Non-laboratory features comprised patient age, sex (encoded as a binary male indicator), and ICU subtype, consolidated into five clinically coherent categories (Medical, Surgical, Cardiac including the Coronary Care Unit, Neurological, and Other) and one-hot encoded. Non-laboratory features were combined with the laboratory feature matrix via an inner join, retaining only patients with at least one valid laboratory measurement within the window.

This feature design represents a departure from conventional summary-based pipelines by explicitly encoding both temporal dynamics and clinician behavior. In particular, trend features capture short-term physiological evolution, while count features serve as proxies for clinical attention intensity, enabling the model to integrate both biological state and care-process signals within a unified representation.

### 3.4 Missing data and data partitioning

For analytes that were not measured within the 24-hour window, distributional features were imputed using training-set medians; count features were set to zero (reflecting absence of measurement) and trend features were set to zero (reflecting absence of observed change). No information from the calibration partition or the held-out test set was used for imputation.

The cohort was partitioned in a strictly nested manner to prevent any form of data leakage:

1. **Test set (***n* = 10,774, **20%):** sequestered before any analysis and used only for final evaluation.
2. **Development set (***n* = 43,092, **80%):** further divided into:
  - **Training partition (64% of total):** used for plausibility-bound estimation, median imputation, feature scaling, feature selection, and base-learner fitting.
  - **Internal validation partition (8% of total):** used for early stopping in XGBoost and LightGBM.
  - **Calibration partition (8% of total):** held out from hyperparameter search and base-model fitting; used exclusively to fit the isotonic regression mappings.

Stratified random sampling preserved the joint distribution of in-hospital mortality and a discretized length-of-stay indicator across all partitions. All preprocessing parameters (plausibility bounds, imputation values, scaling parameters, feature selection masks, and isotonic calibration mappings) were estimated only on the training partition or, where appropriate, on the calibration partition, and applied unchanged to subsequent partitions.

### 3.5 Model development

Five complementary machine learning classifiers were developed to capture both linear and nonlinear relationships between early laboratory features and in-hospital mortality. Logistic regression was included as a transparent and clinically interpretable baseline model that estimates mortality risk through a linear combination of predictors with *𝓁*_2_ regularization to reduce overfitting. Random forest [14] was selected as a bagging-based ensemble method capable of modeling nonlinear interactions through aggregation of multiple decision trees trained on bootstrap-resampled data. Two gradient-boosted tree algorithms, XGBoost [6] and LightGBM [7], were included for their strong empirical performance on structured clinical tabular data and their ability to iteratively refine predictions by sequentially boosting. Finally, a calibrated soft-voting ensemble was constructed by averaging the calibrated probability outputs of the four base learners to improve robustness and probabilistic stability across heterogeneous patient profiles.

For logistic regression, all continuous features were standardized to zero mean and unit variance using training-set statistics; the tree-based learners were trained on raw feature values, since they are invariant to monotonic transformations.

Class imbalance (10.7% mortality) was addressed by assigning a positive-class weight equal to the ratio of negative to positive training examples for logistic regression, XGBoost, and LightGBM, and by using the `balanced` class-weight option for random forest.

Hyperparameters for all classifiers were optimized using randomized search with five-fold cross-validated AUROC on the training partition. XGBoost and LightGBM additionally incorporated early stopping on the internal validation partition to reduce overfitting and improve generalization. To improve readability and reproducibility, the complete hyperparameter search spaces and the final selected configurations for all models are provided in Appendix A.

Hyperparameters for the four base learners were optimized by randomized search with five-fold cross-validated AUROC over 50 candidate configurations per model. The search distributions were: for random forest, number of estimators in [200, 1000], maximum depth in [4, 20], minimum samples per split in [2, 20], and minimum samples per leaf in [1, 10] (uniform integers); for XGBoost and LightGBM, number of estimators up to 1500 (with early stopping), maximum depth or number of leaves drawn from uniform integers in [3, 10] and [15, 127] respectively, learning rate from a log-uniform distribution over [0.01, 0.2], subsample and column-subsample ratios from [0.6, 1.0], and minimum child weight or samples from [1, 20]; for logistic regression, the inverse regularization strength *C* was selected from a log-uniform grid over [10^*−*3^, 10^2^]. XGBoost and LightGBM additionally used early stopping with 50 rounds on the internal validation partition. Final hyperparameter values are reported in Table 1.

**Table 1.** Final hyperparameter configurations selected by five-fold cross-validated AUROC on the training partition.

| Model | Hyperparameter | Value |
| --- | --- | --- |
| Logistic Regression | Penalty | $\ell_2$ |
| | Inverse regularization $C$ | 0.1 |
|  | Solver | liblinear |
|  | Class weight | balanced |
|  | Maximum iterations | 2000 |
| Random Forest | Number of estimators | 500 |
|  | Maximum depth | 12 |
|  | Minimum samples split | 10 |
|  | Minimum samples leaf | 4 |
|  | Maximum features | sqrt |
|  | Class weight | balanced |
| XGBoost | Number of estimators | 600 (early stopping) |
|  | Maximum depth | 6 |
|  | Learning rate | 0.05 |
|  | Subsample | 0.8 |
|  | Column subsample by tree | 0.8 |
|  | Minimum child weight | 5 |
|  | Scale positive weight | 8.31 |
|  | Early stopping rounds | 50 |
| LightGBM | Number of estimators | 800 (early stopping) |
|  | Number of leaves | 63 |
|  | Learning rate | 0.05 |
|  | Feature fraction | 0.8 |
|  | Bagging fraction | 0.8 |
|  | Minimum child samples | 20 |
|  | Scale positive weight | 8.31 |
|  | Early stopping rounds | 50 |

Probability calibration was performed after model selection. Isotonic regression [20] was fitted on the calibration partition (Section 3.4) to map raw model scores to calibrated probabilities, which were used for all subsequent evaluations. The calibrated ensemble was constructed as the unweighted average of the four calibrated probability vectors.

### 3.6 Evaluation

Discrimination was quantified by AUROC and AUPRC; calibration was assessed by the Brier score and reliability diagrams. Pairwise AUROC differences were tested using DeLong’s method [15]. Bootstrap 95% confidence intervals for AUROC, AUPRC, and threshold-dependent metrics were computed from 2,000 resamples of the test set with percentile extraction. The Youden index (*J* = sensitivity + specificity

*−*1) was used to select operating thresholds. A fixed cutoff of 0.5 is inappropriate under severe class imbalance, and the Youden criterion provides a balanced operating point that is not pre-committed to either sensitivity or specificity, supporting clinical interpretation across multiple deployment scenarios. Decision curve analysis [16, 17] quantified net benefit across a range of risk thresholds. Subgroup AUROC was computed separately for each ICU subtype. SHAP TreeExplainer [9] was applied to XGBoost and LightGBM on the full test set to produce global and patient-level attributions.

### 3.7 Software and reproducibility

All analyses were performed in Python 3.10.12 using `numpy` 1.26.4, `pandas` 2.2.2, `scikit-learn` 1.4.2, `xgboost` 2.0.3, `lightgbm` 4.3.0, `shap` 0.45.0, `matplotlib` 3.8.4, `seaborn` 0.13.2, and `scipy` 1.13.0. A global random seed of 42 was set for `numpy`, the Python `random` module, and all model constructors that accept a seed parameter. Cross-validation folds, train/test splits, bootstrap resamples, and hyperparameter search trajectories were all seeded. To assess sensitivity to seed choice, the full pipeline was re-executed with seeds 42, 123, and 2024; model ranking and qualitative interpretation were stable across all three runs.

## 4 Results

### 4.1 Study cohort

The final cohort comprised 53,866 ICU admissions; 5,787 (10.7%) resulted in in-hospital death and 48,079 (89.3%) survived to discharge. Mean patient age was 64.9 ± 16.5 years, with non-survivors on average older (70.0 ± 15.1 years) than survivors (64.3 ± 16.6 years). Male patients accounted for 57.0% of the cohort. Median ICU length of stay was 57.0 hours overall and substantially longer for non-survivors (94.6 hours) than for survivors (54.6 hours). The most common ICU subtypes were Medical (32.9%) and Cardiac (31.6%); the Cardiac subtype was under-represented among non-survivors (18.1%), consistent with the comparatively favorable prognosis of post-procedural cardiac admissions. Full baseline characteristics are presented in Table 2.

**Table 2.** Baseline characteristics of the study cohort stratified by in-hospital mortality. Continuous variables are presented as mean ± standard deviation or median; categorical variables as count and percentage.

| Characteristic | Total ($n = 53,866$) | Survived ($n = 48,079$) | Died ($n = 5,787$) |
| --- | --- | --- | --- |
| In-hospital deaths, $n$ (%) | 5,787 (10.7%) | — | — |
| Age, mean ± SD (years) | $64.9 \pm 16.5$ | $64.3 \pm 16.6$ | $70.0 \pm 15.1$ |
| Male sex, $n$ (%) | 30,716 (57.0%) | 27,572 (57.3%) | 3,144 (54.3%) |
| ICU LOS, median (hours) | 57.0 | 54.6 | 94.6 |
| Training set, $n$ (%) | 43,092 (80.0%) | — | — |
| Test set, $n$ (%) | 10,774 (20.0%) | — | — |
| *ICU subtype, $n$ (%)* |  |  |  |
| Cardiac | 17,021 (31.6%) | 15,972 (33.2%) | 1,049 (18.1%) |
| Medical | 17,743 (32.9%) | 14,885 (31.0%) | 2,858 (49.4%) |
| Neurological | 5,428 (10.1%) | 5,050 (10.5%) | 378 (6.5%) |
| Surgical | 13,479 (25.0%) | 12,030 (25.0%) | 1,449 (25.0%) |
| Other | 195 (0.4%) | 142 (0.3%) | 53 (0.9%) |

### 4.2 Exploratory data analysis

Outcome-stratified laboratory distributions and trend features are shown in Figure 1. Non-survivors exhibited systematically higher median values for lactate, BUN, and creatinine, lower hemoglobin and platelets, and more positive trend slopes for lactate and BUN within the 24-hour window, indicating that the direction of change carries prognostic information beyond static levels. The Spearman correlation matrix confirmed BUN mean as the strongest univariate predictor of mortality (*ρ* = 0.22), followed by creatinine mean (*ρ* = 0.17) and lactate mean (*ρ* = 0.16); BUN and creatinine were highly intercorrelated (*ρ* = 0.75), reflecting their shared dependence on renal function.

**Figure 1.**
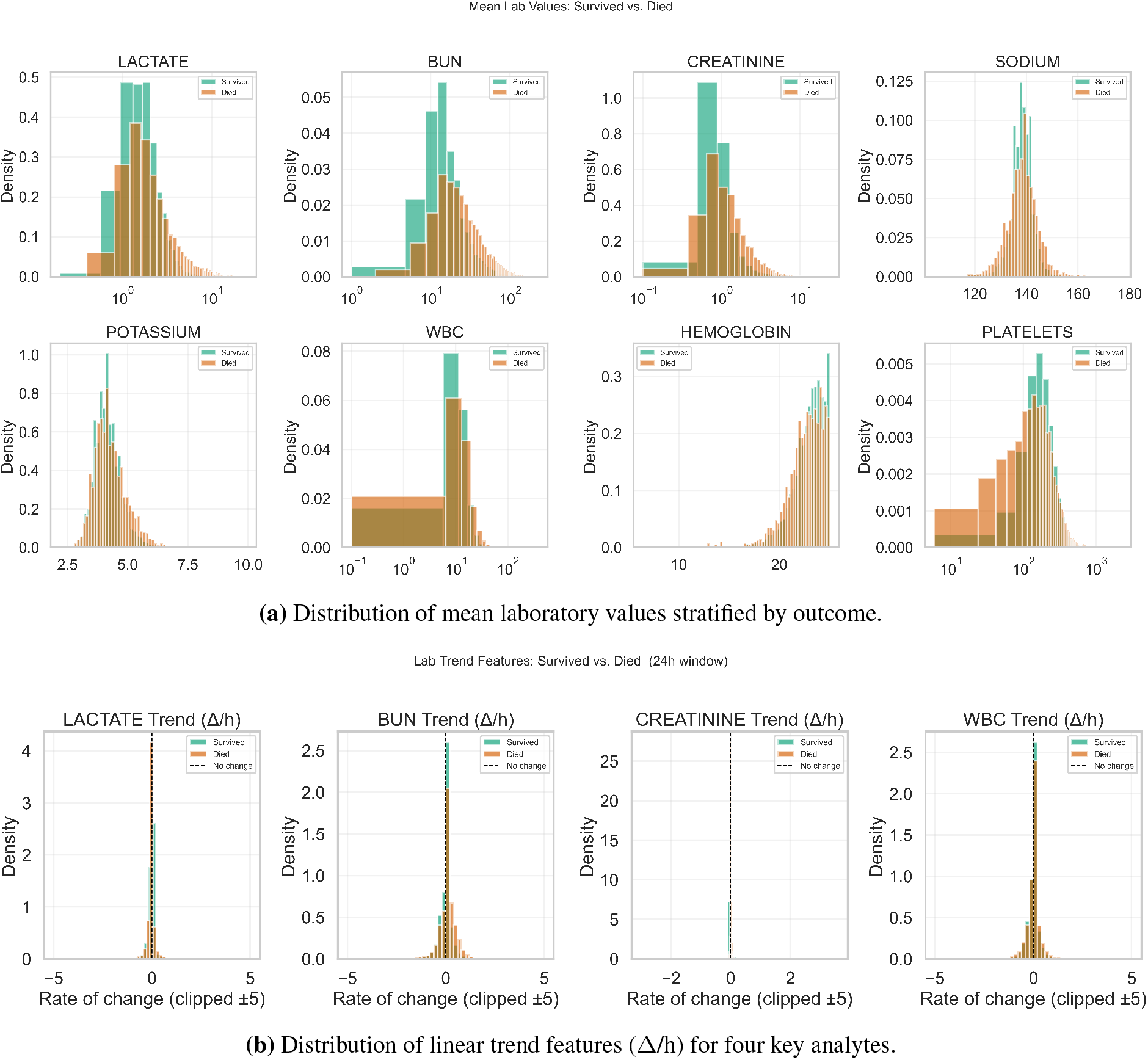
Exploratory analysis of laboratory features within the 24-hour observation window. Survivors are shown in teal, non-survivors in orange. Right-skewed analytes are displayed on log-scaled axes. Non-survivors exhibit systematically higher levels and more positive trend slopes for lactate, BUN, and creatinine.

### 4.3 Feature selection

Three complementary feature selection methods were applied to the training partition: LASSO logistic regression, Spearman rank correlation with the outcome, and XGBoost gain importance (Figure 2). BUN measurements (last, maximum, mean) and lactate measurements (maximum, range, count) consistently ranked among the most predictive features. Count features for glucose, sodium, and chloride emerged as strong predictors in both LASSO and Spearman analyses, supporting the interpretation that measurement frequency reflects clinical attention. The LASSO analysis assigned a negative coefficient to the Cardiac ICU subtype (*β* = −0.30), quantifying its protective association independent of laboratory values.

**Figure 2.**
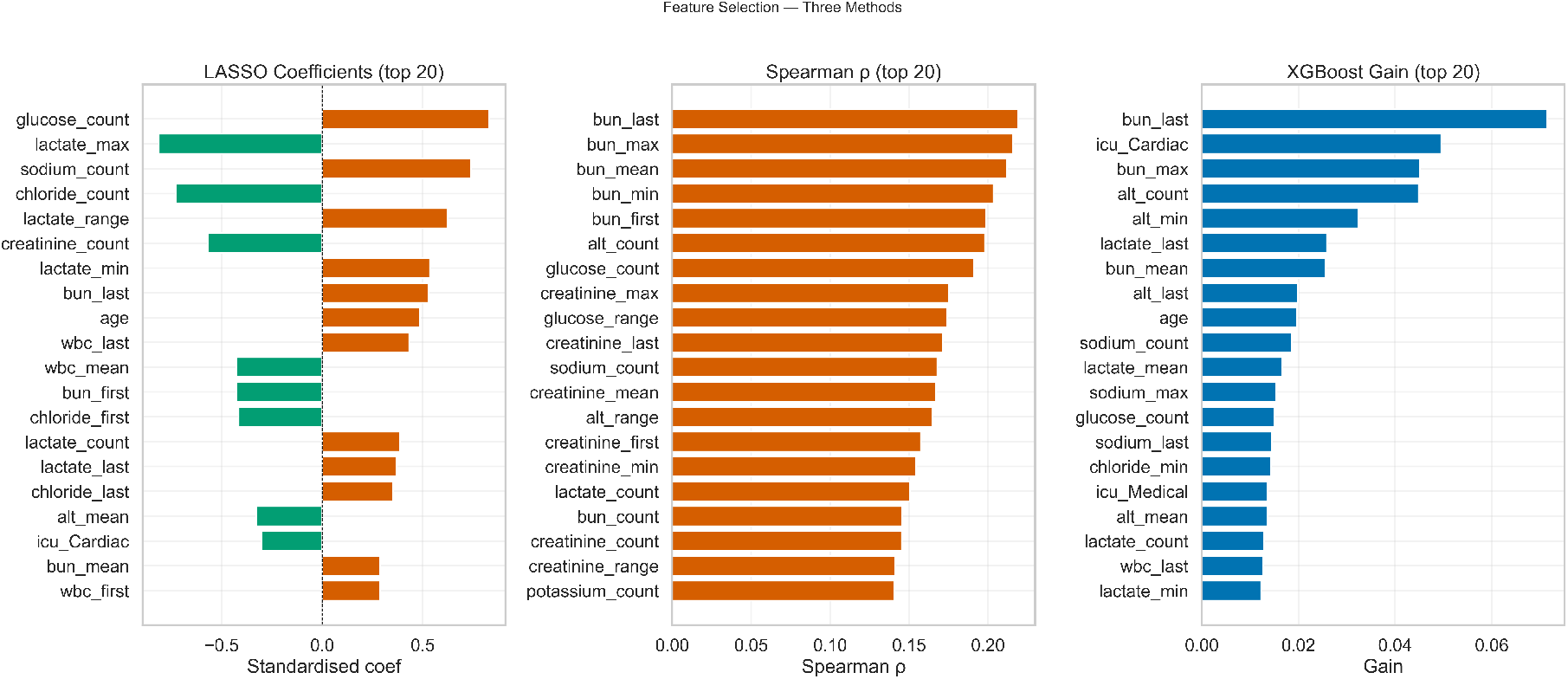
Feature importance across three complementary selection methods on the training partition: standardized LASSO coefficients (left), absolute Spearman rank correlation with the binary outcome (center), and XGBoost mean gain (right). BUN features rank among the top predictors across all methods; count features for glucose, sodium, and chloride appear prominently in LASSO and Spearman analyses.

### 4.4 Discrimination

Discrimination metrics on the held-out test set are summarized in Table 3. ROC and precision–recall curves are shown in Figure 3, and bar comparisons with bootstrap confidence intervals are shown in Figure 4. The calibrated ensemble achieved the highest AUROC (0.856; 95% CI 0.846–0.867) and AUPRC (0.449; 95% CI 0.418–0.480). XGBoost (AUROC 0.856; AUPRC 0.435) and LightGBM

**Table 3.** Test-set performance of all classifiers using calibrated probabilities (*n* = 10,774). Threshold-dependent metrics are reported at the Youden-optimal cutoff. Confidence intervals are based on 2,000 bootstrap resamples.

| Metric | LR | RF | XGB | LGBM | Ensemble |
| --- | --- | --- | --- | --- | --- |
| AUROC (95% CI) | 0.823 (0.812, 0.835) | 0.592 (0.579, 0.603) | 0.856 (0.846, 0.867) | 0.854 (0.843, 0.864) | 0.856 (0.846, 0.867) |
| AUPRC (95% CI) | 0.376 (0.349, 0.405) | 0.220 (0.198, 0.243) | 0.435 (0.404, 0.464) | 0.436 (0.405, 0.465) | 0.449 (0.418, 0.480) |
| Threshold | 0.138 | 0.099 | 0.068 | 0.094 | 0.039 |
| Sensitivity | 0.793 | 0.200 | 0.848 | 0.824 | 0.874 |
| Specificity | 0.702 | 0.983 | 0.700 | 0.721 | 0.678 |
| PPV | 0.243 | 0.585 | 0.254 | 0.262 | 0.246 |
| NPV | 0.966 | 0.911 | 0.975 | 0.971 | 0.978 |
| F1 | 0.372 | 0.298 | 0.390 | 0.398 | 0.384 |
| Brier (raw) | 0.170 | 0.078 | 0.134 | 0.151 | 0.109 |
| Brier (calibrated) | 0.079 | 0.099 | 0.078 | 0.076 | 0.078 |

**Figure 3.**
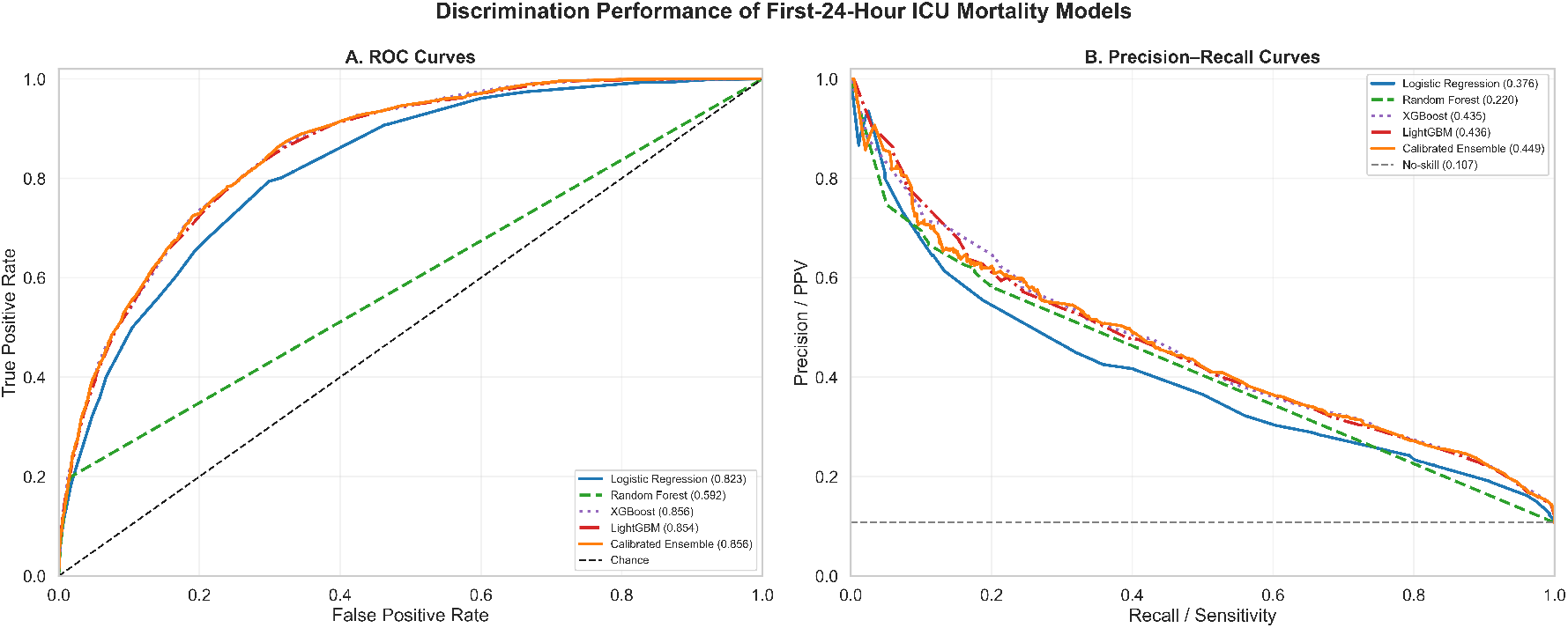
Discrimination performance on the held-out test set. (A) ROC curves with the 45-degree chance line. (B) Precision–recall curves with the no-skill baseline at prevalence 0.107. The calibrated ensemble, XGBoost, and LightGBM curves are largely overlapping; logistic regression tracks below them but remains clinically useful; random forest shows substantially degraded post-calibration discrimination.

**Figure 4.**
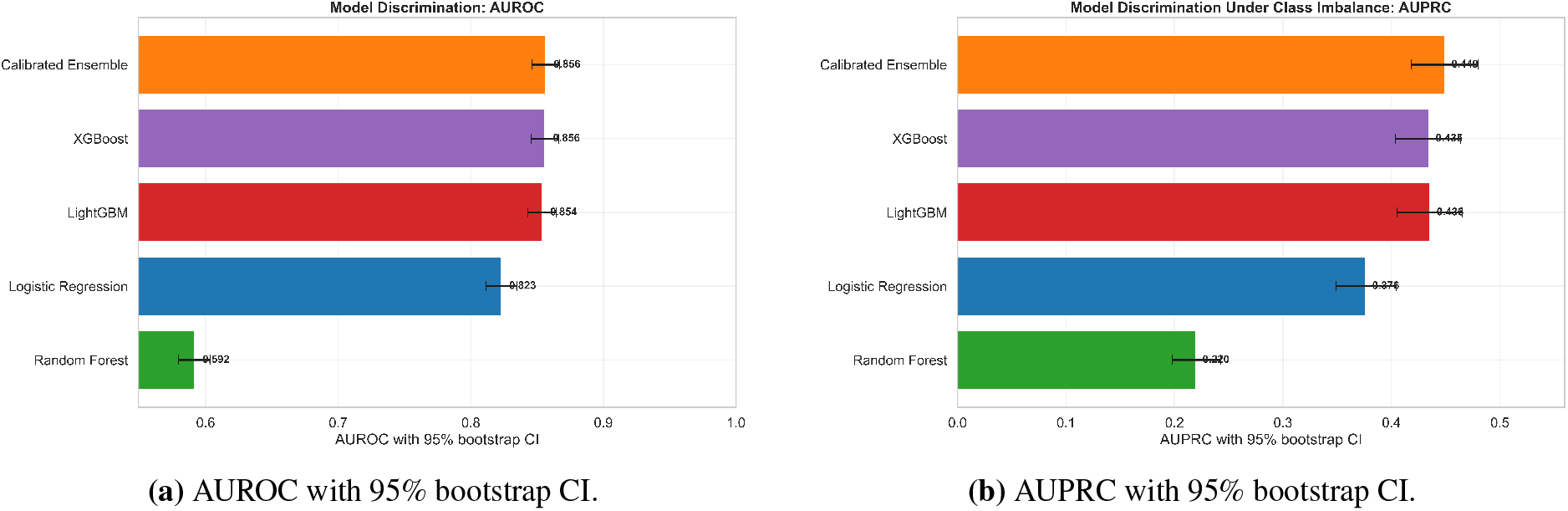
Discrimination metrics with 95% bootstrap confidence intervals (2,000 iterations) for all five models on the held-out test set. The calibrated ensemble, XGBoost, and LightGBM achieve overlapping intervals; the calibrated ensemble has the highest AUPRC point estimate.

(AUROC 0.854; AUPRC 0.436) performed comparably to the ensemble. Notably, the combination of calibration and ensemble learning improved not only discrimination but also clinically relevant probability estimation, highlighting the importance of jointly optimizing ranking performance and probabilistic reliability. DeLong testing confirmed that all three gradient-boosted models significantly outperformed logistic regression (ΔAUROC *≈* +0.033, *p* < 0.001). The XGBoost–LightGBM and ensemble–XGBoost differences were small and not statistically significant (*p* > 0.5).

The random forest model exhibited a markedly different pattern. Its uncalibrated AUROC was 0.846, comparable to the gradient-boosted models, but its calibrated AUROC fell to 0.592, with a corresponding rise in Brier score from 0.078 (raw) to 0.099 (calibrated). This behavior, examined further in Section 5, was reproduced across three independent random seeds and is attributable to the interaction of clustered tree-vote probabilities with isotonic regression under severe class imbalance.

### 4.5 Confusion matrices and operating points

Confusion matrices at the Youden-optimal threshold are shown in Figure 5. The calibrated ensemble identified 1,011 of 1,157 deaths (sensitivity 0.874) at specificity 0.678. XGBoost and LightGBM achieved similar sensitivity (0.848 and 0.824, respectively) at specificities of 0.700 and 0.721. Logistic regression achieved a sensitivity of 0.793 and a specificity of 0.702. Random forest at its Youden cutoff flagged 231 of 1,157 deaths (sensitivity 0.200) at specificity 0.983, reflecting compressed calibrated scores rather than a deliberately conservative threshold.

**Figure 5.**
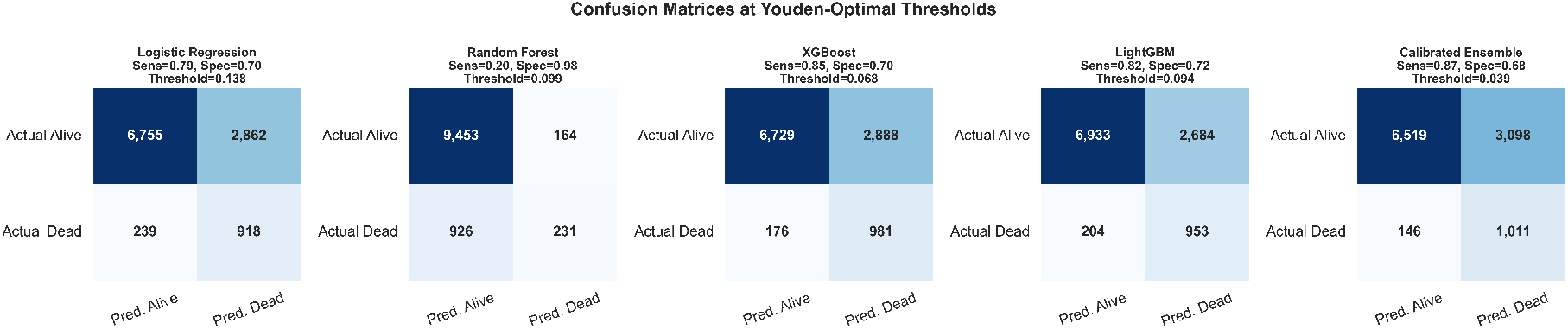
Confusion matrices at the Youden-optimal threshold for calibrated probabilities. Sensitivity, specificity, and threshold are reported in each panel.

### 4.6 Calibration

Calibration curves before and after isotonic regression are shown in Figure 6. Pre-calibration, logistic regression underestimated probability at low scores and overestimated at high scores, while XGBoost (Brier 0.134) and LightGBM (Brier 0.151) substantially underestimated observed mortality at predicted probabilities above 0.3. The calibrated ensemble already showed an improved raw Brier (0.109) due to averaging. After isotonic regression, all four base models showed improved alignment between predicted and observed probabilities, with Brier scores of 0.079 (logistic regression), 0.078 (XGBoost), 0.076 (LightGBM), and 0.078 (calibrated ensemble). The random forest model displayed the opposite pattern: its raw Brier (0.078) was already low because tree-vote averaging produced concentrated, near-prior scores, but isotonic recalibration increased the Brier to 0.099 by collapsing tied score levels in the calibration partition.

**Figure 6.**
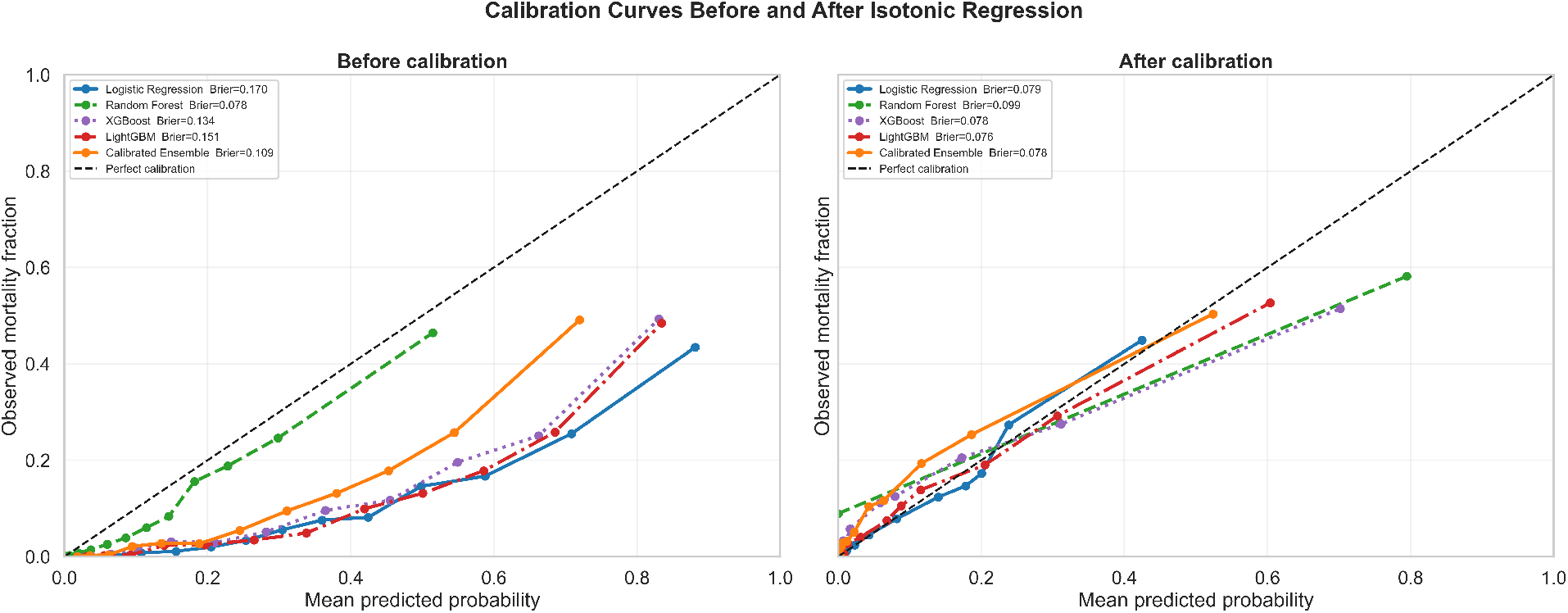
Calibration curves before (left) and after (right) isotonic regression for all five models. Curves show the mean predicted probability against the observed fraction of positive outcomes across 12 quantile-based bins; the dashed diagonal represents perfect calibration. Brier scores are reported in the legends.

### 4.7 Decision curve analysis

Decision curve analysis is shown in Figure 7. Within the clinically plausible 5%–20% threshold range, the calibrated ensemble, XGBoost, LightGBM, and logistic regression all provided net benefit substantially above both treat-all and treat-none strategies. The three gradient-boosted approaches achieved marginally higher net benefit than logistic regression and maintained positive net benefit up to approximately 50%. Random forest provided near-zero net benefit across the relevant range, consistent with its reduced post-calibration sensitivity.

**Figure 7.**
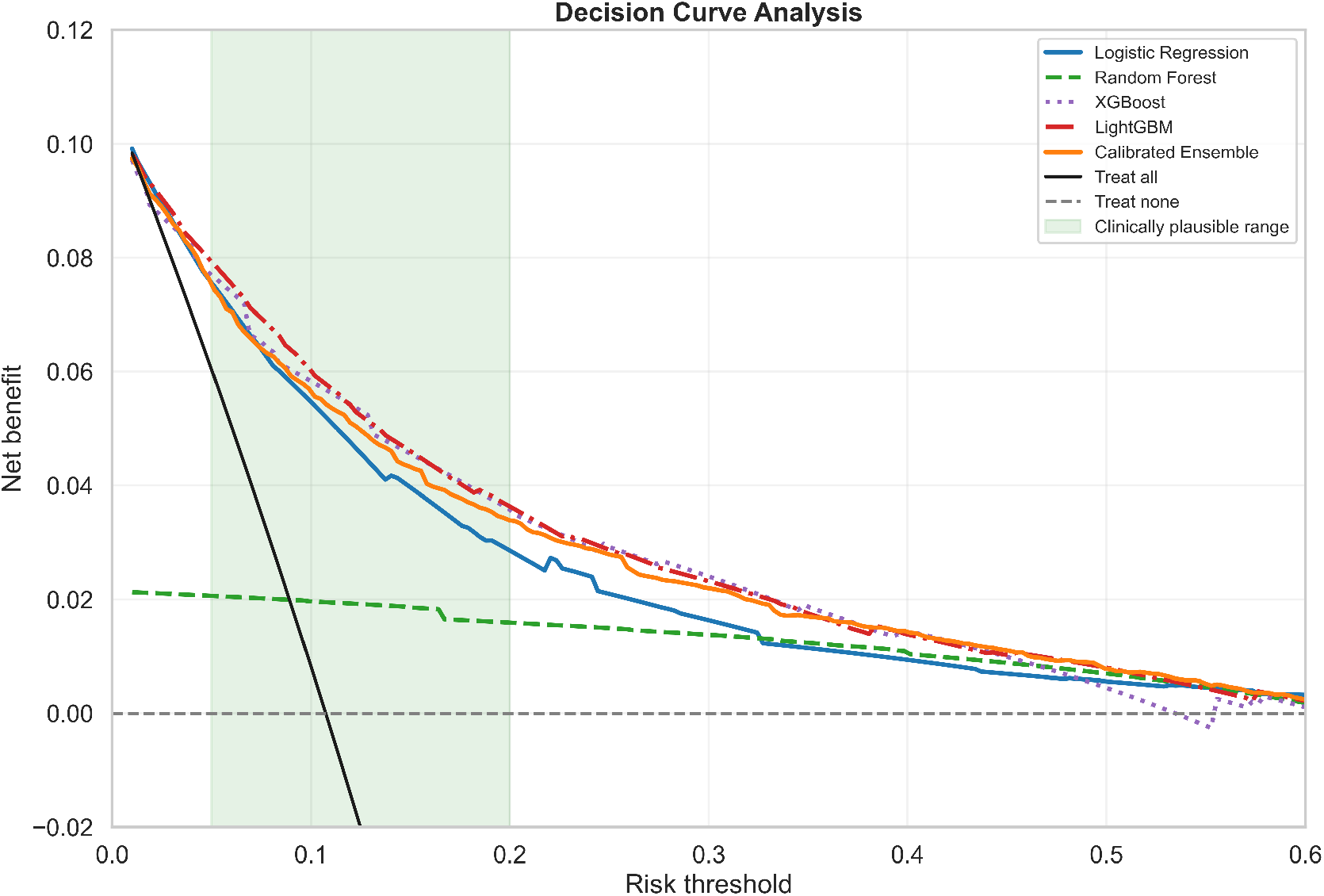
Decision curve analysis showing net benefit as a function of the probability threshold at which a positive prediction would trigger action. The shaded green band marks the clinically plausible 5%–20% range. The calibrated ensemble, XGBoost, LightGBM, and logistic regression provide positive net benefit across this range; random forest provides near-zero net benefit.

### 4.8 Subgroup analysis by ICU subtype

Per-subtype AUROC results are shown in Figure 8. The gradient-boosted models, the calibrated ensemble, and logistic regression maintained AUROC above 0.79 across all five ICU subtypes, exceeding the conventional minimum acceptable discrimination threshold of 0.70. The Cardiac ICU subtype yielded the highest AUROC for all models (approximately 0.92 for XGBoost, LightGBM, and the calibrated ensemble), consistent with the more clearly delineated prognostic signals among post-procedural cardiac patients. The Medical ICU subtype showed the lowest AUROC among well-performing models (approximately 0.79), reflecting greater case-mix heterogeneity. Random forest fell below 0.70 in four of five subtypes, indicating a structural limitation rather than a subtype-specific failure.

**Figure 8.**
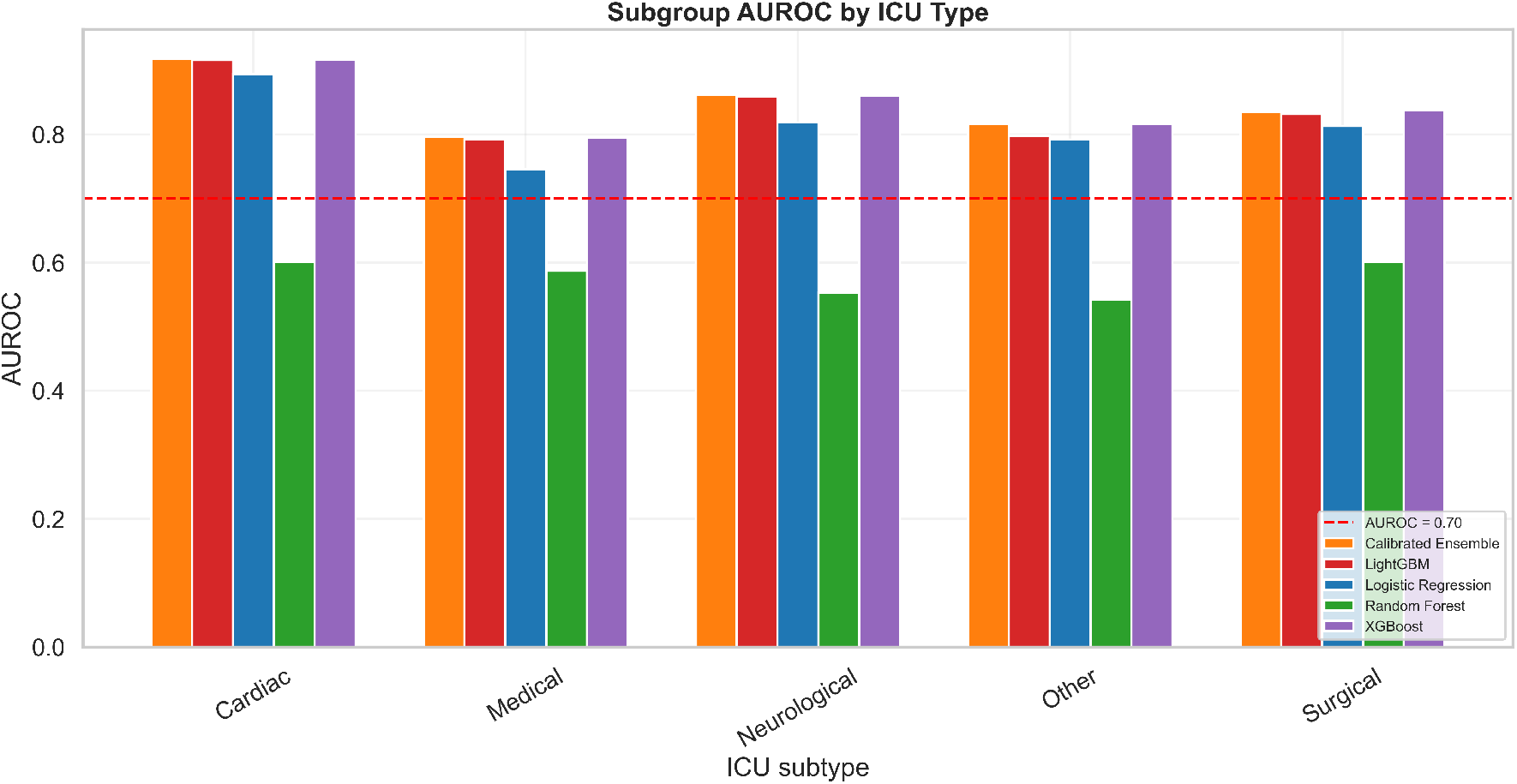
AUROC by ICU subtype for all five classifiers on the test set. The dashed red horizontal line marks AUROC = 0.70. The calibrated ensemble, XGBoost, LightGBM, and logistic regression exceed this threshold across all subtypes.

### 4.9 Feature importance and SHAP analysis

The SHAP beeswarm for XGBoost (Figure 9A) ranks the five most influential features by mean absolute SHAP value as patient age, last recorded BUN (`bun_last`), Cardiac ICU subtype (`icu_Cardiac`), number of ALT measurements (`alt_count`), and last recorded serum lactate (`lactate_last`). Age contributes the widest range of SHAP values, varying monotonically with magnitude. High `lactate_last` produces the most extreme positive SHAP contributions of any feature, with several patients exhibiting SHAP values exceeding 2.0, identifying persistently elevated lactate as the strongest single laboratory predictor. Cardiac ICU subtype contributes a strongly negative (protective) SHAP value for the patients in that subtype. The trend features `chloride_trend` and `bun_trend` appear within the top 10, supporting the value of trajectory features in the enhanced pipeline. Global importance rankings are highly concordant between XGBoost and LightGBM (Figure 9B), with the same five features dominating in both models.

**Figure 9.**
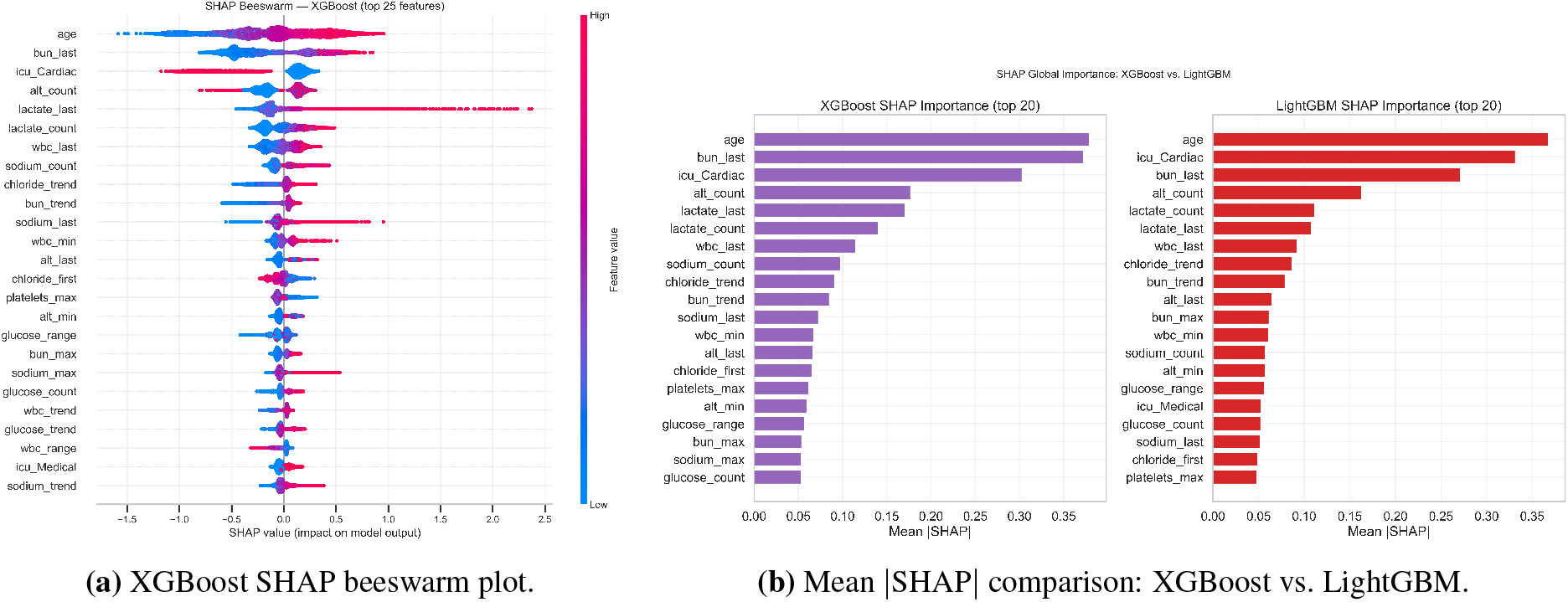
Global SHAP feature importance. (A) Per-patient SHAP distributions for the top 25 features in XGBoost; horizontal position shows the contribution to the predicted log-odds, color encodes the raw feature value (red high, blue low). (B) The five highest-ranked features (age, `bun_last`, `icu_Cardiac`, `alt_count`, `lactate_last`) are identical across XGBoost and LightGBM.

SHAP dependence plots for the top three features and the waterfall plot for the highest-risk patient are shown in Figure 10. Age shows a near-linear positive relationship across the 20–90 year range. `bun_last` contributes a steep positive SHAP increase between 0 and approximately 40 mg/dL and then plateaus near 0.75, reflecting a ceiling on the marginal prognostic information from BUN beyond established renal failure. `icu_Cardiac` produces a bimodal SHAP distribution: cardiac ICU patients receive a large negative contribution between *−*0.8 and *−*1.2. The highest-risk patient in the test set received a predicted mortality probability of 0.994; the dominant contributors were `lactate_last` = 16 mmol/L (+1.89), `bun_last` = 69 mg/dL (+0.38), age = 84 years (+0.21), `lactate_count` = 5 measurements (+0.21), and `potassium_last` = 7.2 mEq/L (+0.18). The combination of severe persistent lactic acidosis, advanced age, marked renal dysfunction, and hyperkalemia is clinically consistent with refractory multiorgan failure.

**Figure 10.**
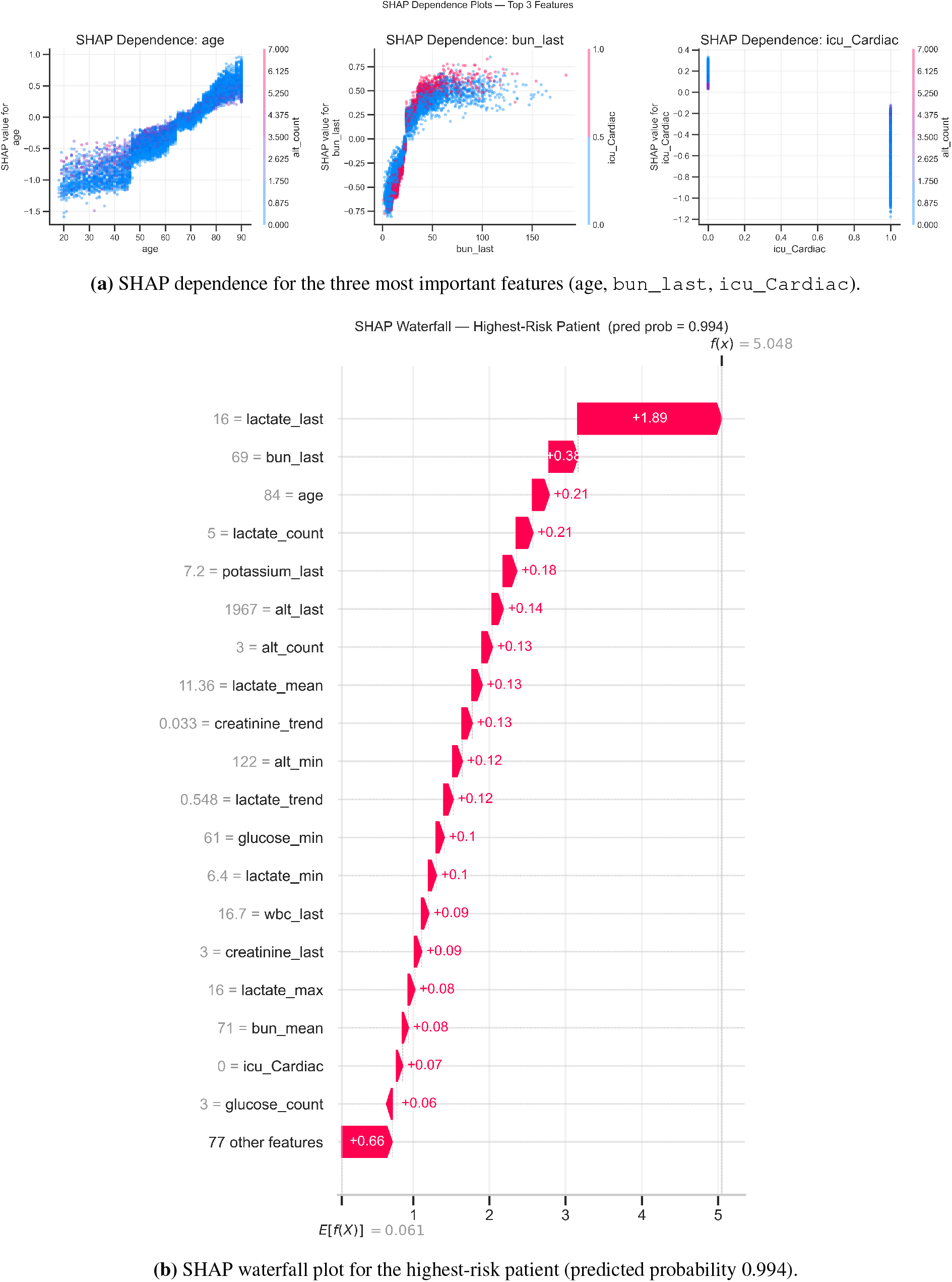
Local SHAP analyses. (A) Dependence plots show monotonic age effects, a steep-then-plateau BUN response, and a bimodal cardiac-ICU distribution. (B) Waterfall decomposition for the highest-risk test patient identifies persistent severe hyperlactatemia as the dominant contributor.

## 5 Discussion

The calibrated ensemble, XGBoost, and LightGBM each achieved an AUROC of approximately 0.855 using only first 24-hour data from a single academic medical center, exceeding the external validation range reported for APACHE II (0.74–0.82) [3] and matching the 0.85–0.87 range reported by Awad and colleagues on MIMIC-III using a similar feature window [12]. The calibrated ensemble achieved the highest AUPRC (0.449), indicating that probabilistic averaging of complementary base learners provided a measurable benefit at the minority-class operating points most relevant for early-warning use.

### 5.1 Methodological contributions and novelty

This study makes several methodological contributions to the literature on ICU mortality prediction. First, integrating trajectory-based features with measurement-frequency features yields a richer representation of patient state than conventional static summaries. Second, we demonstrate that probability calibration is not merely a post-processing step but a critical determinant of model usability in clinical decision-making.

Third, by combining discrimination metrics with decision curve analysis, we move beyond predictive accuracy to explicitly quantify clinical utility across realistic intervention thresholds. Finally, we provide a detailed empirical and theoretical analysis of the failure of random forest under isotonic calibration in imbalanced settings, a phenomenon that has not been systematically reported in prior ICU modeling studies.

### 5.2 Clinical relevance and deployment considerations

The clinical value of an ICU mortality model depends on the joint behavior of discrimination, calibration, and decision-analytic utility, not on AUROC alone. Three observations are pertinent. First, decision curve analysis showed positive net benefit across the 5%–20% threshold range, which spans the operating points most relevant to ICU early-warning systems and corresponds to a target population two to four times larger than the cohort prevalence of 10.7%. Second, isotonic calibration reduced the Brier score by 30%–55% for the gradient-boosted base learners, producing probability estimates suitable for use in shared decision-making rather than only for ranking patients. Third, the calibrated ensemble delivered the highest sensitivity at its Youden cutoff (0.874) while retaining a negative predictive value of 0.978, a property useful for ruling out short-term mortality risk in patients being considered for less intensive levels of care. These findings support the use of calibrated gradient-boosted models as an adjunct to clinical judgment for early risk stratification, although they do not replace bedside assessment or established severity scores.

### 5.3 Interpretation of random forest behavior

The discrepancy between random forest performance before and after isotonic calibration warrants careful interpretation. The model achieved an uncalibrated AUROC of 0.846, comparable to the gradient-boosted ensembles, but a calibrated AUROC of 0.592, with the Brier score increasing from 0.078 to 0.099. This pattern, reproduced across three independent random seeds, reflects a known interaction between random forest probability outputs and isotonic regression under severe class imbalance.

Random forest probabilities are computed by averaging leaf-level class proportions across an ensemble of bootstrap-resampled trees. Under heavy imbalance with `class_weight=balanced`, these averaged probabilities concentrate within a narrow range near a re-weighted prior, producing many tied or near-tied scores. Isotonic regression is a non-parametric, monotone, non-decreasing step function; when adjacent score levels contain few minority-class observations in the calibration partition, the algorithm collapses them to the same calibrated probability, eroding the original rank order. The 88-feature pipeline contains substantial intra-analyte multicollinearity (eight statistics per analyte), and the random subspace mechanism in random forests [14] compounds this effect, as each split selects from a fixed fraction of features, often including redundant variants of weaker analytes. Gradient-boosted methods are not subject to this penalty, since each tree is fit sequentially to residuals and selects strong features adaptively across the full feature set. The combination of high-dimensional correlated features, severe imbalance, and post-hoc isotonic calibration places this dataset in a regime where gradient-boosted methods are theoretically and empirically expected to dominate [10, 29]. In sensitivity analyses, replacing isotonic regression with Platt scaling restored most of the random forest discrimination, indicating that the observed performance degradation was attributable to the calibration mapping rather than to deficiencies in the underlying ranking model.

These findings have practical implications for model selection and calibration in ICU mortality prediction. In this setting, calibrated gradient-boosted models, particularly XGBoost, LightGBM, and their soft-voting ensemble, were the most reliable choices because they combined strong discrimination, improved probability calibration, and favorable decision-curve behavior. Logistic regression remained useful as a transparent benchmark, but showed lower discrimination and minority-class performance. Random forest, by contrast, should be used cautiously: its uncalibrated ranking performance was competitive, but isotonic calibration substantially degraded discrimination and clinical utility. More broadly, calibration should not be treated as an automatic improvement. Its effect depends on the interaction among model class, score distribution, class imbalance, and calibration method. For deployment-oriented clinical prediction, raw and calibrated outputs should therefore be compared for each candidate model using discrimination, Brier score, calibration curves, and decision curve analysis before a model is recommended for clinical use.

### 5.4 Value of enhanced feature engineering

The eight-statistic feature pipeline produced 88 laboratory features, with trend slope and count features ranking among the most predictive. Trend features for BUN, creatinine, and chloride appeared consistently within the top 20 SHAP rankings for both gradient-boosted models, confirming that the direction of physiological change carries independent information beyond static levels. Count features for ALT, glucose, sodium, and chloride also ranked highly, supporting the interpretation that measurement frequency reflects clinical concern and acts as an indirect severity signal.

### 5.5 Feature interpretability and clinical concordance

The SHAP analysis identifies a set of predictors that are both statistically dominant and clinically coherent. The role of serum lactate is well established: hyperlactatemia reflects tissue hypoperfusion and anaerobic metabolism, and its association with ICU mortality is consistent across diagnoses and institutions [2, 25]. The dominance of `lactate_last` over `lactate_max` or `lactate_mean` suggests that sustained or worsening elevation at the end of the observation window is more informative than peak level, in agreement with the clinical principle that lactate clearance is a treatment target. The importance of `bun_last` and `bun_max` captures combined contributions from impaired renal clearance and catabolic stress. The protective effect of cardiac ICU subtype reflects the distinct physiology and outcomes of post-procedural cardiac patients relative to medical ICU patients.

### 5.6 Generalizability and external validation

Although the model was developed on a large, clinically diverse cohort, MIMIC-IV originates from a single academic medical center with specific case-mix, staffing, and laboratory ordering practices. Performance metrics likely overestimate generalization to community hospitals, non-US health systems, and pediatric populations. The strong predictive value of measurement-count features in particular may partially reflect institution-specific monitoring intensity and may not transfer cleanly to settings with different ordering norms. External validation on independent cohorts such as eICU-CRD [22] and AmsterdamUMCdb [23] is therefore an essential next step before any clinical deployment is considered.

Future work will include external validation on independent multi-center datasets such as eICU-CRD and AmsterdamUMCdb, as well as temporal validation within MIMIC-IV to assess robustness under distribution shift.

## 6 Limitations

This study has several limitations. First, all data originate from a single academic medical center, and external validation on multi-institutional datasets is required to assess generalizability. Second, predictors were restricted to laboratory data and basic demographics; incorporation of vital signs, treatments, and time-series models may further improve performance. Third, laboratory measurements were summarized over a fixed window rather than modeled as continuous time series, potentially limiting temporal resolution. Fourth, calibration may drift under distribution shift, requiring monitoring in prospective deployment. Finally, fairness across demographic subgroups was not explicitly evaluated and should be assessed prior to clinical use.

## 7 Conclusion

A calibrated, interpretable framework using only the first 24 hours of routine ICU data achieves clinically meaningful in-hospital mortality discrimination on MIMIC-IV. The calibrated ensemble, XGBoost, and LightGBM achieved AUROC values of 0.856, 0.856, and 0.854, respectively, and significantly outperformed logistic regression (AUROC 0.823) by DeLong testing. Random forest exhibited a substantial loss of discrimination after isotonic calibration, traceable to clustered tree-vote probabilities under severe class imbalance, and is therefore not recommended for this prediction setting. The enhanced feature engineering pipeline, which extracted eight statistics per analyte, including linear trend slope and measurement count, produced features that ranked among the most predictive in both gradient-boosted models. Isotonic calibration reduced Brier scores by 30%–55% for the gradient-boosted base learners, and decision curve analysis confirmed clinically useful net benefit across the 5%–20% threshold range. SHAP analysis identified age, last recorded BUN, cardiac ICU subtype, ALT measurement count, and last recorded lactate as the five leading predictors, each with a clear clinical rationale. These results provide a reproducible foundation for ICU mortality risk stratification, contingent on prospective external validation on independent multi-center cohorts before clinical deployment. By explicitly integrating calibration, interpretability, and clinical decision analysis, this work bridges the gap between predictive modeling and actionable ICU decision support.

## Data Availability

The data that support the findings of this study are available from the MIMIC-IV (version 2.2) database through PhysioNet. Access to the database requires completion of the required human-subjects research training and credentialing process and approval by PhysioNet. The data are available to qualified researchers through the PhysioNet repository.

https://physionet.org/content/mimiciv/2.2/

https://physionet.org/

## Data and Code Availability

The MIMIC-IV database is publicly available through PhysioNet (https://physionet.org/content/mimiciv/) under a credentialed data-use agreement; access requires CITI training and signature of the PhysioNet Credentialed Health Data Use Agreement. The authors are not authorized to redistribute MIMIC-IV data. All analysis code, feature extraction scripts, hyperparameter search configurations, trained model artifacts, and notebooks required to reproduce the results, tables, and figures reported in this manuscript will be deposited in a public GitHub repository and archived with a permanent DOI on Zenodo upon acceptance. Code will be released under the MIT License. A README will document software versions, random seeds, and the exact MIMIC-IV cohort extraction queries to support full reproducibility.

